# Impact of vaccine hesitancy on secondary COVID-19 outbreaks in the US

**DOI:** 10.1101/2021.09.21.21263915

**Authors:** Alfonso de Miguel Arribas, Alberto Aleta, Yamir Moreno

## Abstract

The COVID-19 outbreak has become the worst pandemic in at least a century. To fight this disease, a global effort led to the development of several vaccines at an unprecedented rate. There have been, however, several logistic issues with its deployment, from their production and transport, to the hesitancy of the population to be vaccinated. For different reasons, an important amount of individuals is reluctant to get the vaccine, something that hinders our ability to control and - eventually - eradicate the disease. In this work, we analyze the impact that this hesitancy might have in a context in which a more transmissible SARS-CoV-2 variant of concern spreads through a partially vaccinated population. We use age-stratified data from surveys on vaccination acceptance, together with age-contact matrices to inform an age-structured SIR model set in the US. Our results clearly show that higher vaccine hesitancy ratios led to larger outbreaks. A closer inspection of the stratified infection rates also reveals the important role played by the youngest groups. Our results could shed some light on the role that hesitancy will play in the near future and inform policy-makers and the general public of the importance of reducing it.

## I. INTRODUCTION

More than a year and a half since the onset of the COVID-19 outbreak, firstly reported by the Chinese authorities on December 31, 2019, it is clear that this pandemic has become the worst one in at least a century.

Multiple aspects of our life have been severely affected at various scales: psychological [1, 2] and social [3]; human-related systems and infra-structures [4, 5]; supply chains [6, 7]; and the economy in general [8, 9]. To manage the disease, it was mandatory to adopt a plethora of measures aimed at reducing the mixing and interaction among individuals in order to mitigate SARS-CoV-2 transmission and propagation. Lockdowns [10–14], curfews and mobility restrictions [15–18], social distancing [19–21], personal prophylaxis [22, 23] are now part of the new normalcy across the world. This “new normal” [24–26], being its impact as critical as the virus itself, was conceived and promised as something that should be ephemeral, a toll to pay, just until the ultimate solution arrives: the vaccines.

A rapid and massive scientific effort [27, 28] to develop a vaccine against SARS-CoV-2 was deployed and successfully achieved in less than a year; another unprecedented fact. From December 2020, just a year after the onset of the pandemic, several nations started their vaccination campaigns in the pursue of herd immunity to control the pandemic. But again, further problems proliferate. From lack of confidence due to the relative short time for vaccine development and approval, to the typical fears of suffering serious side effects or due outlandish conspiracy theories, some people hesitate or are reluctant to vaccination [29–32]. For good or bad, this phenomenon is neither exclusive nor new [29, 33]. To be vaccinated (or not) is, in most countries, a choice of the individual, even though the consequences of such a choice go beyond the self and affect the social sphere. Hesitancy poses an ethical problem since if a critical fraction of individuals declines vaccine uptake for any disease, resurgence is to be expected [34–36]. This already happened in the UK, which was declared measles-free in 2017 but lost this status just 2 years later due to sub-optimal vaccination uptake[37].

In this work, we explore, via modeling, the consequences of vaccine hesitancy in successive waves of COVID-19. To this end, we focus on the US. In particular, given the large heterogeneity in vaccine hesitancy across the US, we perform our analysis on each state, although our analysis and conclusions could be extrapolated elsewhere provided relevant data is at disposal. We make use of an age-structured SIR model to simulate the spreading dynamics, which is fed with real and up-to-date data of the US age-distributed population and contact matrices, as well as with survey-based seroprevalence estimations [38]. Despite these elements of realism, it is worth noting that we do not intend to replicate the real trajectory of the COVID-19 pandemic in the United States until now, neither we aim to accurately forecast the unfolding of future outbreaks and epidemic sizes. Rather, we try to answer the following question: what would happen if a certain fraction of the population hesitates or declines vaccine uptakes and normalcy is re-established (as it is now the case in many places, not only the US) or new aggressive variants emerge? We propose a scenario in which COVID-19 outbreaks emerge in each state, independently, with a mitigated propagation due to the presence of some restrictions, while there is an ongoing vaccination campaign designed following the information obtained from public surveys [39]. Once the vaccination and this first outbreak end, we assume a *back to normal* situation, where all restrictions are lifted and disease awareness vanishes, and a new outbreak sets in. To make our simulations more realistic, we assume that these successive outbreaks happen for a more transmissible variant of the virus, mimicking in this way the evolutionary path of the SARS-CoV-2 variants of concern that have evolved towards more transmissible forms of the virus. Our results show that higher vaccine hesitancy rates result in increasing sizes of potential subsequent outbreaks. However, this correlation does not always translate into more deaths, as mortality depends on how non-vaccinated individuals are distributed across age groups and their mixing. Altogether, our results confirm that either the fraction of non-vaccinated individuals or the remaining proportion of susceptible individuals could be good predictors of the size of secondary outbreaks.

## II. MATERIAL AND METHODS

### A. Epidemic model

Given the utmost relevance of age in the effects of COVID-19, it is compulsory to introduce the age distribution of the population and the specific interaction between age groups to adequately model the dynamics of the disease. We use the estimated age-contact matrices provided by Mistry et al. [40] updated to the population structure of 2019 [41]. Then, we build an age-structured SIR model defined by this set of equations[40]:

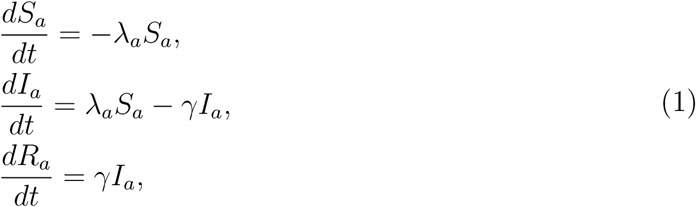

where *S*_*a*_ is the number of susceptible individuals of age *a, I*_*a*_ is the number of infected individuals of age *a, R*_*a*_ is the number of removed individuals of age *a*, and *γ*^*−*1^ is the infectious period, which is assumed to be the same for all age classes and equal to *γ*^*−*1^ = 4.5 days. COVID-19 is a disease with a more complex natural history than a SIR model can account for, being required to add some pre-symptomatic or asymptomatic compartments, as well as a latency period, for certain applications. Nonetheless, it has been shown that SIR models can correctly describe the overall evolution of the disease [42], which is enough for the scope of this paper. Lastly, *λ*_*a*_ is the force of infection for individuals of age *a* and it is expressed as

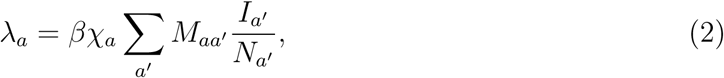

where *β* is the transmissibility of the virus, *N*_*a*_ is the total number of individuals of age *a*, and *M*_*aa*_′ measures the average number of contacts of an individual of age *a* with individuals of age *a*′. Finally, *χ*_*a*_ is an age-dependent susceptibility factor accounting for the lower susceptibility of children to the disease, i.e. *χ*_*a*_ = 0.56 if *a* ≤ 19 and 1 otherwise [43].

The basic reproductive number *R*_0_ is defined in this model as

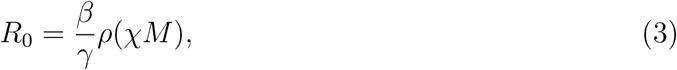

where *ρ*(*χM*) is the spectral radius, or largest eigenvalue, of the age-contact matrix (in this case also incorporating the susceptibility factor) [44].

### B. Scenario

First, we collect the seroprevalence data measured in September 2020 for each age group and US state. We set the corresponding fraction of the population in each state into the removed compartment. Second, we simulate an initial outbreak with an *R*_0_ = 1.5. This basic reproductive number is below the estimated *R*_0_ value for unmitigated transmission of the original variant of SARS-CoV-2 that is around 2.5-3 [45]. With this choice, we mimic a scenario in which there are some restrictions, social distancing and other prophylactic measures in place, yielding a smaller effective *R*_0_. During this outbreak, we implement a vaccination campaign (described below). At the end of the campaign, all individuals that have not refused vaccination shall be vaccinated.

Once the vaccination campaign is completed, we assume that societies have returned to normalcy, i.e., any kind of restrictions and precautionary measures are lifted. Then, a new outbreak is seeded in each state, emulating a spill over from other states in the country or importation from other countries. In this second outbreak, we set *R*_0_ = 6 which is closer to the currently dominant variant of concern (delta variant) [46]. Note that we assume that no awareness or other non-pharmaceutical interventions are in place during this outbreak. Thus, it can be thought as the worst case scenario of resurgence after a vaccination campaign.

As a visual example of the proposed scenario, in figure 1 we show how the incidence would evolve at the level of state for the full epidemics if no vaccination campaign were deployed during the first outbreak. When an aggressive variant sets in, secondary outbreaks may still cause havoc. The inset depicts the evolution of the prevalence, which can reach almost the whole population for large enough *R*_0_. In section 3, we explore how the vaccination efforts modify this baseline scenario.

**FIG. 1.**
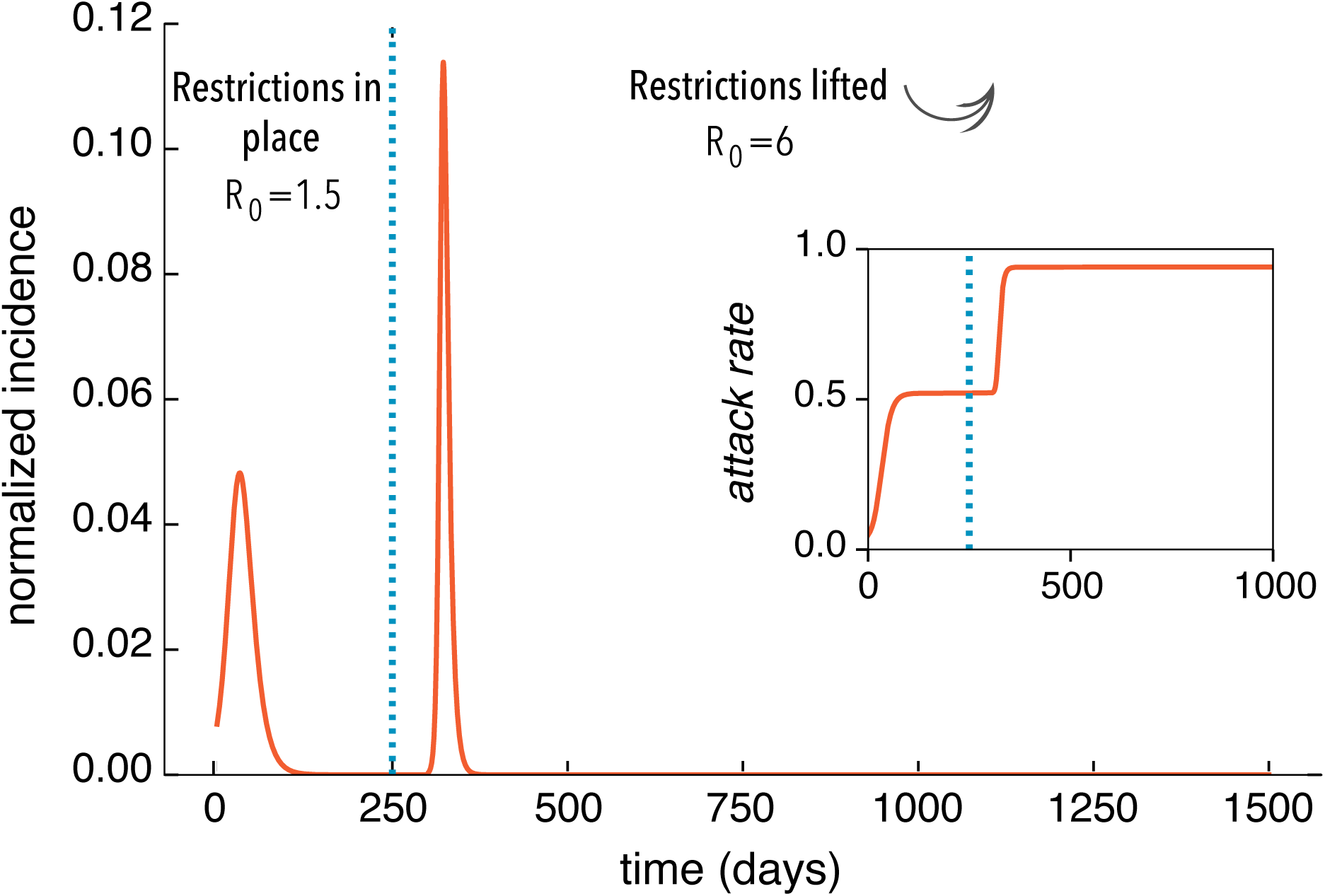
Proposed baseline scenario: following the first wave of the epidemic, part of the population acquires natural immunity. Then, we simulate the propagation of a mitigated outbreak due to the presence of some restrictions, social distancing and prophylaxis measures, leading to a slower propagation of the original variant of the disease (*R*_0_ = 1.5). After the outbreak extinguishes a back-to-normal situation is assumed and all prevention measures are lifted. Then, an outbreak is seeded again with a higher basic reproductive number, *R*_0_ = 6. On top of this baseline scenario, we will introduce a vaccination campaign during the first outbreak and explore the impact of vaccination hesitancy on the second outbreak.

### C. Vaccination

We use data from The COVID States Project (https://covidstates.org), in particular the surveys in Report #43: COVID-19 vaccine rates and attitudes among Americans [39]. These surveys provide information on vaccination acceptance/hesitancy by age at the state level. Therein, several degrees of predisposition toward vaccines are reported. The following categories are distinguished: individuals who are “already vaccinated”, individuals who are inclined to be vaccinated “as soon as possible”, “after at least some people I know”, “after most people I know”, and finally people who “would not get the COVID-19 vaccine”. The shares of people in each category is given at a national level for different age groups. The data shows an important amount of heterogeneity in each of those categories by age group. However, at the level of state, the data is not disaggregated by age groups, only the share of people in each vaccine acceptance category is shown. More specifically, we are looking for the coefficients 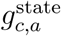, which represent the share of people for every acceptance category, *c*, is the population of the and age-class, *a* in every US state. These coefficients satisfy 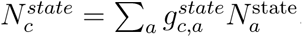, Where 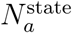 is the population of the state in the age class *a* and 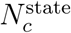, state in the acceptance category *c*. These 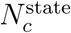 values are provided in appendix A within the report, but the information is not disaggregated by age at the level of state [39].

The report offers information at a national level about how people are distributed within acceptance categories by age groups. We refer to the shares shown in the report as 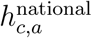, which are normalized by age-class, that is, 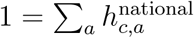 for a particular category *c*. The quantity 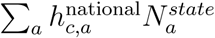 would be the number of people if national coefficients apply for a certain state and vaccine acceptance category *c*. We relate these coefficients 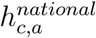 to coefficients 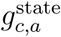 through a linear transformation:

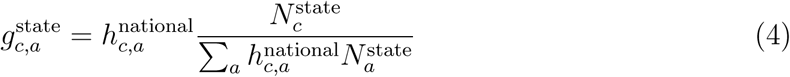

This transformation preserves the shares of people in a certain vaccine acceptance category c in every state and also allows for the introduction of age heterogeneity adapted from the national-level data.

Vaccination campaigns are complex and depend highly on several properties of the population: age, risk groups, professions, supplies, infrastructure, etc. Since we are mostly interested in the aftermath after the vaccination campaign, we adopt a simple scheme. From the aforementioned surveys, we extract the fraction of the population within each age group and state that is willing to be vaccinated, 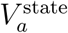. We set the length of the vaccination campaign to be Δ*t*_*v*_ = 150 days and assume that the fraction of population vaccinated per unit time is constant and equals to 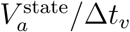. Both susceptible and recovered individuals can be vaccinated. For simplicity, the vaccine is assumed to be 100% effective in preventing the infection.

## III. RESULTS AND DISCUSSION

In figure 2, we show how the incidence and prevalence of the disease changes from the baseline scenario depicted in 1 when vaccination is in place. In particular, we consider the state with the highest vaccine hesitancy, Oklahoma (OK), and the state with the lowest one, Massachusetts (MA). Additionally, for a fairer comparison, the simulations were started with a null initial condition for prevalence (*R*_*a*_(*t* = 0) = 0) (i.e. considering that the whole population is in the susceptible state). Dotted lines in the figure show the case without vaccination. We can see that the impact, in each isolated outbreak and for the full epidemic, is more or less the same for both states, differences owing to population internal structure. When vaccination is introduced in the model (continuous lines), we can appreciate the reduction of peak incidence and epidemic final sizes for both states during the first outbreak. However, when we simulate the second outbreak, the state with the lowest vaccine hesitancy shows a remarkably lower impact, while the other state experiences a sizable second outbreak. The peak of the outbreaks is similar in both outbreaks for Massachussetts, while in Oklahoma, the secondary outbreak is around twice as large as the first outbreak.

**FIG. 2.**
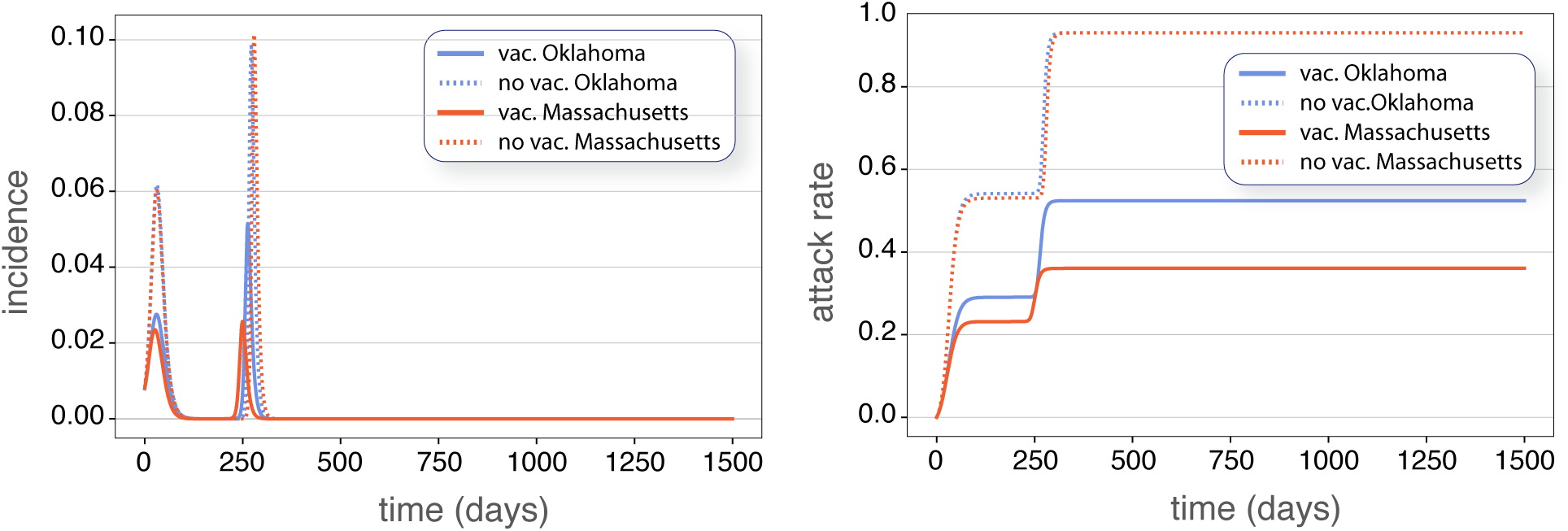
Comparison of peak incidences and final epidemic sizes for the states of Oklahoma (OK), which has the highest vaccine hesitancy, and Massachusetts (MA), where the vaccine hesitancy is the lowest according to surveys [39]. Continuous trajectories (blue and red) represent the simulation with vaccination campaign, whereas dotted trajectories represent the simulation without introducing the vaccination campaign. All simulations started with a fully susceptible population.

Next, we focus on the overall effect of vaccination on the spreading. We explore the relationship between the final attack rate (total fraction of the population that was infected) and the fraction of non-vaccinated individuals in each state. Currently, underage people are not being vaccinated. Thus, this set of individuals is composed by both underage people and adults who manifested vaccine hesitancy in the aforementioned surveys [39].

In order to look for a possible correlation between state-level attack rates and the fraction of non-vaccinated individuals, we performed a linear regression. Figure 3 (A) shows a scatter plot of the attack rates versus the fraction of non-vaccinated individuals for the simulated full epidemic unfolding in every state. The correlation coefficient, *R*^2^ = 0.936 shows a clear relationship between attack rates and vaccine hesitancy for the full period. Note that we have added a simulation for the whole country (the red dot in the scatter plots) with the age-structure from the whole population. Figure 3 (B) shows a scatter plot of the attack rate of the second outbreak, versus the fraction of remaining susceptible individuals at the end of the first outbreak. Here, the correlation coefficient is also very high, *R*^2^ = 0.971. Note that the use of the remaining susceptible fraction rather than directly the fraction of non-vaccinated individuals owes to the fact that once the first outbreak and the vaccination campaign have ended, the demographic structure of the pool of susceptible individuals has changed dramatically. This pool is all conformed by individuals that either declined vaccination or are underage. Since, according to data, hesitancy rates are low in older people, there is a predominance now of younger susceptible individuals. Additionally, figure 4 shows the very same data of figure 3 (A) on the map of the United States. We observe some geographical clustering, even though we are treating each state as a completely isolated population. The states with higher attack rates or, similarly, the states with a higher fraction of vaccine hesitancy, are concentrated mainly in the interior of the country (inner Pacific west, Intermountain, ranging from north (Midwest) to south (inner Southeast).

**FIG. 3.**
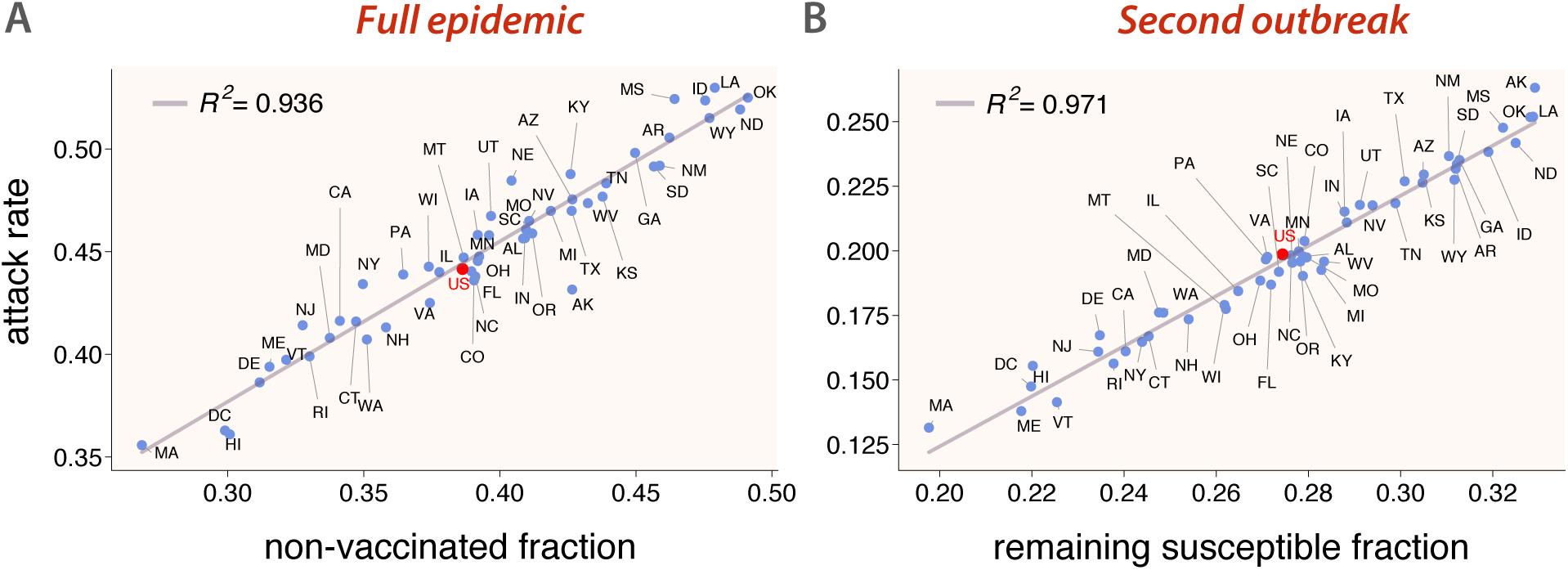
Scatter plot of attack rates after the full epidemic (first outbreak with *R*_0_ = 1.5 and the second one with *R*_0_ = 6) versus the non-vaccinated fraction of individuals (A), and attack rates of the second outbreak (*R*_0_ = 6) versus the remaining susceptible fraction after the first outbreak (B) for every US state. The red dot corresponds a simulation on a population representing the whole country. It is clearly seen that higher hesitancy translates into higher attack rates.

**FIG. 4.**
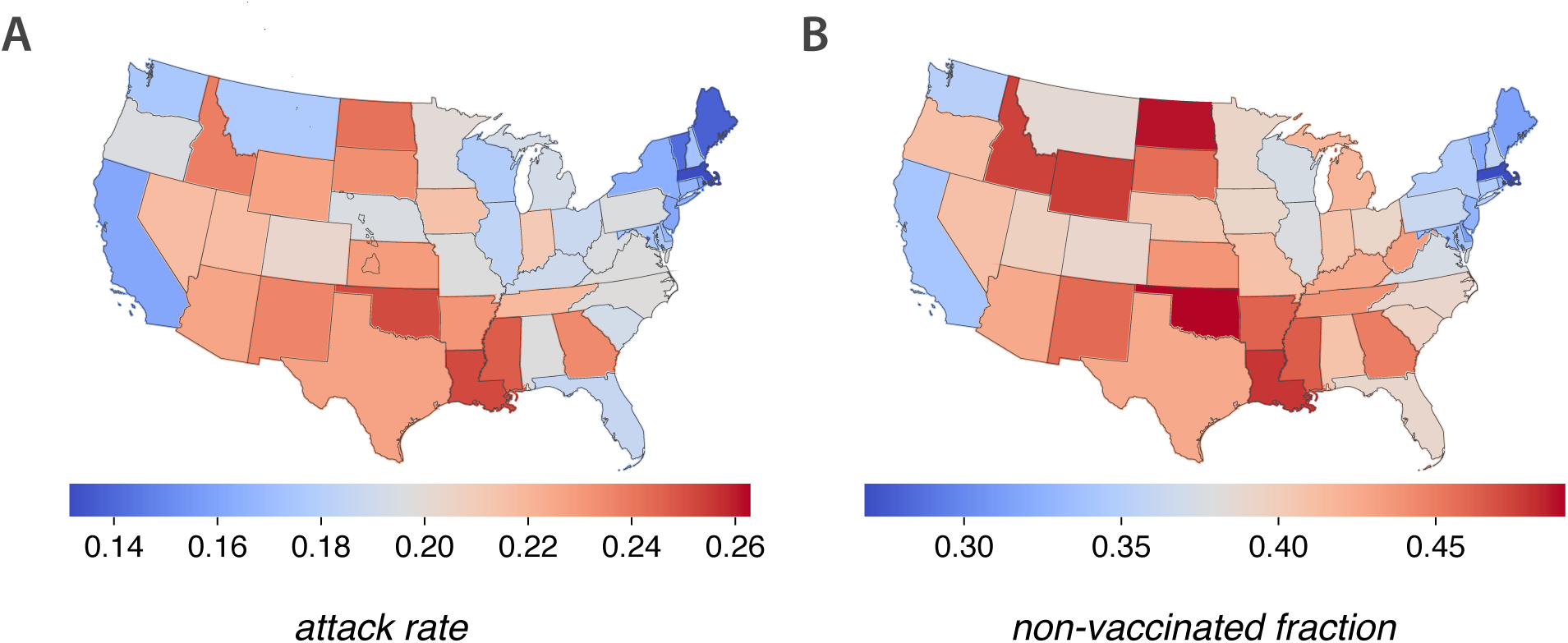
Representation on the US map of the attack rates of every state after the end of the epidemic trajectory proposed in this paper (A), and the fraction of non-vaccinated individuals (B). Some spatial clustering can be appreciated along the country, even though in the simulations all states are completely isolated.

Let us next try to get a deeper understanding of what is happening during the second outbreak. Looking at some particular extreme examples, we can appreciate that the state of Massachusetts (MA), with the lowest vaccine hesitancy (9% of adult population), has the lowest epidemic size during the first outbreak and also during the second outbreak. On the other hand, Alaska (AK) shows one of the lowest attack rates in the first outbreak, but the highest one in the second outbreak, together with the highest fraction of remaining susceptible at the end of the first outbreak, whereas its hesitancy amounts to 23% of the adult population, way behind the most reluctant states. Interestingly, there are other states with a relative low hesitancy rate that also show a sizable second outbreak. This is the case of the state of Utah (UT), with a hesitancy of about 15% among the adult population but nevertheless ranking high in the size of the second outbreak. One could hypothesize that these two states should have a similar number of deaths during the second outbreak. But, remarkably, as we show below, there is indeed more than a simple extrapolation of the correlation between the outbreak size and the number of non-vaccinated/susceptible individuals when it comes to forecast mortality. The reason is that the age of non-vaccinated and/or remaining susceptible matters, not only because it usually determines behavior (and risk of infection) but also because the infection fatality rate heavily depends on it.

In figure 5, we show a scatter plot of deaths per million individuals in the second outbreak versus the fraction of non-vaccinated individuals at the end of the first outbreak. We estimate the number of deaths in each age group by applying the corresponding infection fatality rate (IFR) [47], so that:

**FIG. 5.**
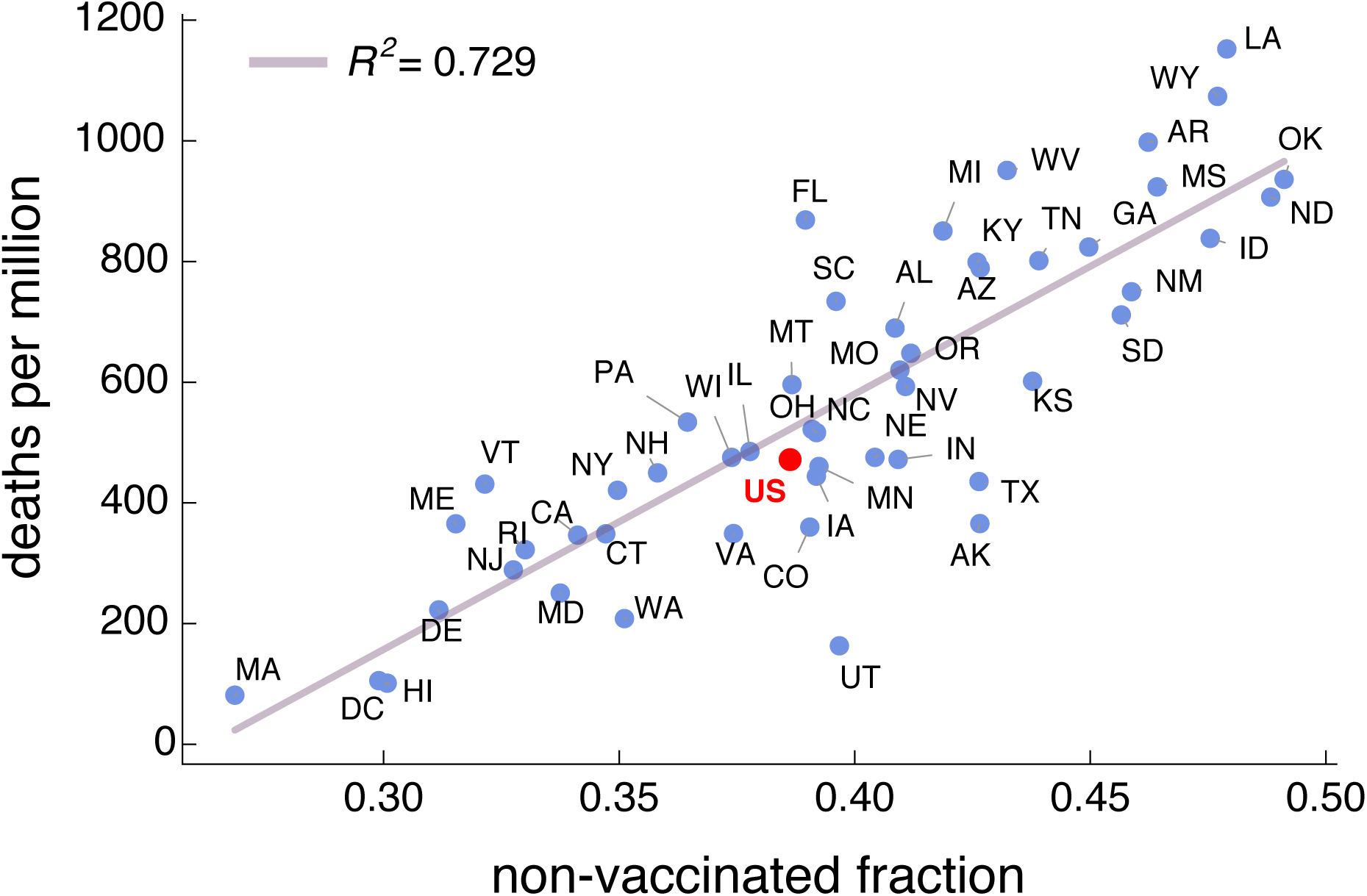
Scatter plot of deaths per million during the second outbreak versus the non-vaccinated fraction at the end of the first outbreak for every US state. Results also shown for a simulation of the epidemic for the whole country as if it were a single age-structured population (red dot). The model does not include deaths as part of the dynamics, but they can be estimated by applying the infection fatality rate to the final fraction of individuals in the removed compartment for each age class (eq. (5)).

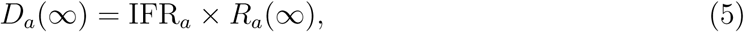

where *R*_*a*_(∞) and *D*_*a*_(∞) are, respectively, the prevalence and the number of deceased individuals at the end of a particular outbreak.

Even if a high correlation coefficient is obtained, its explanatory power is smaller than for the attack rate, which suggests that there are other factors playing a relevant role. Certainly, we can appreciate that higher proportions of deaths tend to occur in those states with higher hesitancy. Bringing back the case of Alaska (AK), and contrary to what could be naively expected, we see that it has been overtaken by several states. Even more striking is the case of Utah (UT), being in the lower part of the ranking. This reveals that apart from vaccine hesitancy, the age structure plays a key role in the disease dynamics and COVID-related fatalities.

To understand better these interdependencies, we next look at the attack rates during the second outbreak by coarse-graining age groups. In figure 6, we show results for 0-18 (A), 18-45 (B), 45-65 (C), and more than 65 year old age groups (D). For each one, the attack rates during the second outbreak are computed as *R*_*a*_(∞)*/R*(∞), while the fractions of remaining susceptible individuals at the end of the first outbreak are computed as *S*_1*a*_*/S*_1_, where *R*(∞) is the final attack rate, and *S*_1_ is the total fraction of remaining susceptible subjects. Thus, these figures tell us the share of people in each group *a* within the susceptible and removed pools.

**FIG. 6.**
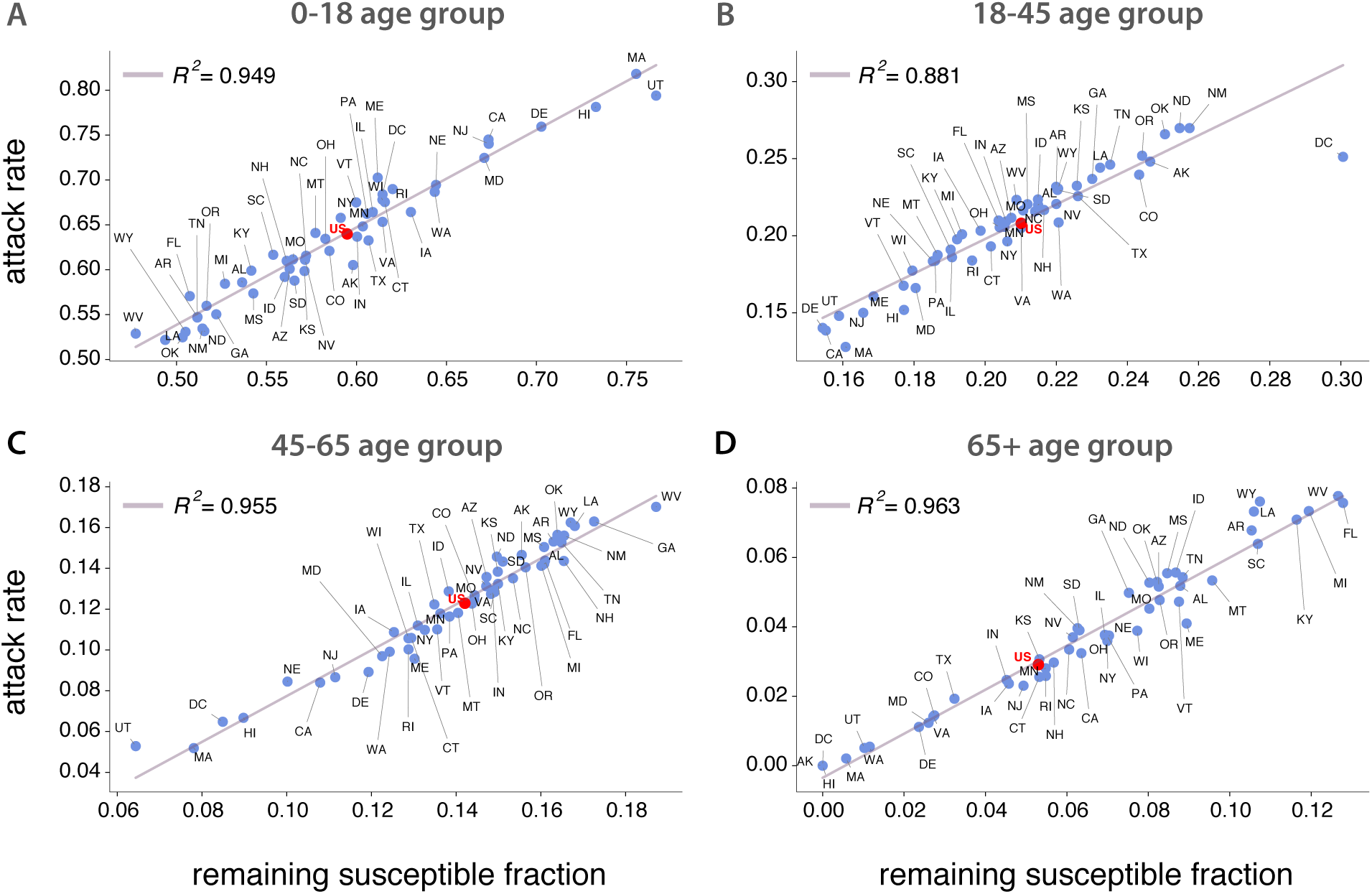
Scatter plot of attack rates during the second outbreak versus the remaining susceptible fraction for every US state. Top-left (A): 0-18 years old group. Top-right (B): 18-45 years old group. Bottom-left (C): 45-65 years old group. Bottom-right (D): over 65 years old group. Results also shown for a simulation of the epidemic for the whole country as if it were a single age-structured population (red dot). These high correlations show also the relevant role of age structure in the disease propagation.

The results by age groups exhibit a very high correlation for the linear fittings, which indicate the relevance of age structure in the transmission of the disease. For every age group, states with higher hesitancy tend to experience larger epidemic sizes. Regarding the cases mentioned before, namely, Utah (UT) and Alaska (AK), one can see that their fractions of remaining susceptible individuals are large in the youngest age groups and rather small (null for Alaska) in the 65+ age strata. This explains why these two states undergo large second outbreaks that are not translated into a higher number of deaths. Finally, we also note that the fraction of remaining susceptible individuals is the highest for roughly every state in the two youngest age brackets (around or higher than 50%), which means that the younger age groups will be the driving group of the second outbreak.

Finally, we look for an estimation of how many deaths could potentially be averted just by reducing the fraction of individuals in the “would not get the COVID-19 vaccine” category in one percentage point. It may occur that for states with an important share of younger population and not very high hesitancy, an extra effort does not pay off. Conversely, in states with an older population and for those with high hesitancy, such additional increase in the percentage of vaccinated may represent an important benefit. Table I shows the number of averted deaths per million people if vaccine hesitancy is reduced by one per cent during the vaccination campaign in every state. A look at the table reveals that, in general, states with the higher number of averted deaths during the second outbreak would be those ranking highest in the fraction of non-vaccinated individuals. This is, as explained before, mediated by the age of the population that remains susceptible and not vaccinated.

**TABLE I.** Average number of averted deaths per million (95% CI in brackets), separately in the 1st and 2nd outbreak, due to reducing vaccine hesitation in one percent point. Results shown for every state and for the whole country (US).

| State | Averted deaths<br>(1st outbreak) | Averted deaths<br>(2nd outbreak) | State | Averted deaths<br>(1st outbreak) | Averted deaths<br>(2nd outbreak) |
| --- | --- | --- | --- | --- | --- |
| AK | 5.12 [2.83- 10.57] | 16.78 [9.37- 35.14] | MT | 12.03 [6.62- 23.86] | 34.49 [18.93- 67.57] |
| AL | 8.62 [4.73- 17.13] | 39.18 [21.49- 76.94] | NC | 8.08 [4.44- 16.11] | 34.3 [18.83- 67.7] |
| AR | 10.23 [5.61- 20.29] | 42.38 [23.17- 82.64] | ND | 10.83 [5.92- 21.52] | 35.56 [19.34- 69.23] |
| AZ | 9.66 [5.29- 19.12] | 39.74 [21.73- 77.58] | NE | 9.75 [5.34- 19.35] | 27.7 [15.11- 54.23] |
| CA | 8.79 [4.81- 17.45] | 24.96 [13.62- 49.08] | NH | 12.1 [6.67- 24.09] | 33.04 [18.19- 65.35] |
| CO | 7.66 [4.21- 15.43] | 26.57 [14.62- 53.06] | NJ | 4.49 [2.47- 8.91] | 27.55 [15.08- 54.42] |
| CT | 10.43 [5.72- 20.71] | 28.18 [15.43- 55.57] | NM | 10.73 [5.9- 21.38] | 37.28 [20.45- 73.19] |
| DC | 4.09 [2.22- 8.28] | 9.43 [5.16- 20.01] | NV | 7.57 [4.16- 15.1] | 34.65 [19.04- 68.39] |
| DE | 8.34 [4.59- 16.6] | 26.71 [14.71- 52.98] | NY | 4.18 [2.29- 8.29] | 31.78 [17.35- 62.54] |
| FL | 10.3 [5.62- 20.2] | 48.74 [26.54- 94.48] | OH | 10.55 [5.79- 20.98] | 32.53 [17.83- 63.96] |
| GA | 5.89 [3.24- 11.8] | 38.9 [21.38- 76.83] | OK | 9.74 [5.34- 19.41] | 38.24 [20.93- 74.83] |
| HI | 9.31 [5.09- 18.44] | 10.87 [6.06- 22.76] | OR | 11.05 [6.07- 21.97] | 35.75 [19.61- 70.23] |
| IA | 7.31 [4.0- 14.56] | 31.54 [17.25- 62.05] | PA | 7.84 [4.3- 15.2] | 36.33 [19.88- 71.2] |
| ID | 10.18 [5.59- 20.29] | 35.19 [19.29- 68.9] | RI | 10.26 [5.63- 20.38] | 26.66 [14.59- 52.55] |
| IL | 9.28 [5.08- 18.44] | 30.61 [16.73- 60.12] | SC | 9.66 [5.32- 19.19] | 39.87 [21.91- 78.17] |
| IN | 9.00 [4.95- 18.04] | 30.06 [16.51- 59.43] | SD | 10.53 [5.79- 21.03] | 35.13 [19.24- 68.87] |
| KS | 9.08 [4.99- 18.16] | 33.38 [18.29- 65.69] | TN | 9.54 [5.24- 19.0] | 39.14 [21.47- 76.84] |
| KY | 11.88 [6.52- 23.52] | 37.12 [20.32- 72.48] | TX | 5.99 [3.29- 12.05] | 27.89 [15.3- 55.49] |
| LA | 7.68 [4.22- 15.25] | 45.27 [24.79- 88.51] | UT | 5.68 [7.38- 26.53] | 17.02 [9.31- 34.05] |
| MA | 8.37 [4.59- 16.67] | 16.94 [9.29- 33.83] | VA | 8.31 [4.57- 16.66] | 27.93 [15.37- 55.55] |
| MD | 5.64 [3.1- 11.27] | 25.95 [14.27- 51.71] | VT | 13.40 [7.38- 26.53] | 31.36 [17.21- 61.56] |
| ME | 14.13 [7.77- 27.89] | 29.7 [16.29- 58.21] | WA | 8.14 [4.48- 16.35] | 22.54 [12.4- 45.11] |
| MI | 12.03 [6.6- 23.82] | 40.84 [22.34- 79.66] | WI | 10.70 [5.88- 21.28] | 30.5 [16.71- 59.9] |
| MN | 7.42 [4.08- 14.82] | 31.34 [17.18- 61.9] | WV | 13.25 [7.28- 26.2] | 44.72 [24.5- 87.12] |
| MO | 10.60 [5.81- 21.04] | 34.9 [19.09- 68.37] | WY | 12.39 [6.82- 24.64] | 39.78 [21.81- 77.73] |
| MS | 8.78 [4.82- 17.48] | 38.48 [21.09- 75.36] | US | 7.95 [4.36- 15.85] | 32.15 [17.62- 63.43] |

## IV. CONCLUSIONS

In this work, we have explored SARS-CoV-2 transmission dynamics on a population that is partially vaccinated and is seeded again with the virus when restrictions are fully lifted. We explored, in particular, to what extent vaccine hesitancy may still drive sizable outbreaks in a context where a more transmissible SARS-CoV-2 variant of concern is dominant. We used data from vaccination acceptance surveys, together with up-to-date age distributed populations and contact matrices in the US to inform an age-structured SIR model. Our results show a clear correlation between the size of experienced outbreaks, once all kinds of measures are lifted, and the fraction of vaccine hesitancy or, similarly, the fraction of remaining susceptible individuals at the onset of a second outbreak. Higher vaccine hesitancy ratios expose the population to larger outbreaks and, inversely, higher vaccine acceptance ratios can mitigate the impact to the point of negligible secondary waves due to immunity of the population. We have also inspected in detail the role of the age structure of the population in both the attack rate and the mortality of secondary outbreaks. Our findings reveal that the prevalence is highly correlated with the fraction of remaining susceptible individuals by age classes, with the youngest contributing the most to the attack rate. It is however not immediate to project such a correlation to the expected number of deaths, as here too age plays a role, though in the opposite direction, e.g., the younger the population, the lower the mortality. Finally, we estimated the number of potentially averted deaths during the course of the simulated epidemic if the number of people reluctant to vaccine uptake were reduced in one percentage point. Results again clearly show that states with the higher fraction of vaccine hesitancy can potentially avert a higher number of deaths.

Summarizing, our model is able to quantify the effects of vaccine hesitancy in the dynamics of secondary waves of COVID-19. The most important implications of the results reported here include: *(i)* data on vaccination by age is important to accurately capture the evolution of mortality in secondary waves; *(ii)* allocation of additional resources is more important in states with relative high hesitancy rates but specially in states where the remaining susceptible population is older; *(iii)* reintroduction of restrictions could be needed in states with very high attack rates to reduce pressure over healthcare systems; and *(iv)* incentives to vaccination directed towards the younger population will reduce the prevalence, while they will reduce the number of deaths if they focus on the older generations.

Finally, we acknowledge that our model has several limitations. One is at the core of its compartmental structure, not including a more detailed progression of the natural history of the disease, which might affect our estimation of deaths, and does not consider hospitalizations of any kind. The vaccination campaign could be implemented in a more realistic way and owing to each state idiosyncrasy but, more importantly, vaccines are revealing to be not sterilizing and thus not fully preventing transmission and, on top of that, immunity decays with time. These facts do not affect the overall dynamics explored in this paper, but should be incorporated to provide reliable estimations on the exact amount of expected infections or deaths. Additionally, the behavioral responses are not completely accounted for. All these factors open important challenges for future works. What if COVID-19 cannot be fully eradicated with the current generation of vaccines? What levels of endemicity could our healthcare systems cope with in the mid-term? Finally, our analysis represents hypothetical scenarios that can unveil mechanisms and correlations rather than producing accurate forecasts. Yet, these scenarios can be helpful for policy making and providing quantitative arguments to the public debate about the role of vaccination in mitigating and containing disease propagation.

## Data Availability

This article has no additional data.

## Notes

### Competing Interest Statement

The authors have declared no competing interest.

### Funding Statement

A. dM., A. A and Y.M. acknowledge partial support from the Government of Aragon and FEDER funds, Spain through grant E36-20R (FENOL), and by MINECO and FEDER funds (FIS2017-87519-P). A. dM is funded by an FPI Predoctoral Fellowship of MINECO. A.A. and Y.M. acknowledge support from Banco Santander (Santander-UZ 2020/0274) and the financial support of Soremartec S.A. and Soremartec Italia, Ferrero Group. The funders had no role in study design, data collection, and analysis, decision to publish, or preparation of the manuscript.

### Author Declarations

No IRB is needed for this research study

